# Estimation of risk factors for COVID-19 mortality - preliminary results

**DOI:** 10.1101/2020.02.24.20027268

**Authors:** F Caramelo, N Ferreira, B Oliveiros

## Abstract

Since late December 2019 a new epidemic outbreak has emerged from Whuhan, China. Rapidly the new coronavirus has spread worldwide. China CDC has reported results of a descriptive exploratory analysis of all cases diagnosed until the 11th February 2020, presenting the epidemiologic curves and geo-temporal spread of COVID-19 along with case fatality rate according to some baseline characteristics, such as age, gender and several well-established high prevalence comorbidities. Despite this, we intend to increase even further the predictive value of that manuscript by presenting the odds ratio for mortality due to COVID-19 adjusted for the presence of those comorbidities and baseline characteristics such as age and gender. Besides, we present a way to determine the risk of each particular patient, given his characteristics.

We found that age is the variable that presents higher risk of COVID-19 mortality, where 60 or older patients have an OR = 18.8161 (CI95%[7.1997; 41.5517]). Regarding comorbidities, cardiovascular disease appears to be the riskiest (OR= 12.8328 CI95%[10.2736; 15.8643], along with chronic respiratory disease (OR=7.7925 CI95%[5.5446; 10.4319]). Males are more likely to die from COVID-19 (OR=1.8518 (CI95%[1.5996; 2.1270]). Some limitations such as the lack of information about the correct prevalence of gender per age or about comorbidities per age and gender or the assumption of independence between risk factors are expected to have a small impact on results. A final point of paramount importance is that the equation presented here can be used to determine the probability of dying from COVID-19 for a particular patient, given its age interval, gender and comorbidities associated.

## Introduction

At the end of December of 2019, a cluster of pneumonia cases in Wuhan has created an alert worldwide. These cases were thought to be associated with a seafood market, which was closed on the 1^st^ of January 2020. At that time there were 41 confirmed cases and one death, which occurred in a patient with serious underlying medical conditions thus it was not clear to attribute that death to the illness. The alert from China CDC had already been made to the World Health Organization^1-3^. Potential causes for that cluster of disease, such as influenza, adenovirus, severe acute respiratory syndrome (SARS-CoV) and Middle-East respiratory syndrome (MERS-CoV) were expunged.^1,3-4^ In fact, the pathogen causing this severe illness was quickly identified as a novel coronavirus and its genomic characterization was rapidly performed enabling to create a suitable test method^1-5^. The genomic characterization showed some relations both to SARS-CoV and MERS-CoV^4-6^, whilst it was thought that this new strand would be less aggressive than those two coronavirus ^4-6^. Nevertheless, on the 20th of January the onset changed due to the confirmation of human-to-human transmission ^2,7-10^. The first death attributed to this novel coronavirus, named as SARS-CoV-2 (COVID-19 is the associated disease), occurred on January 13, and according to the China CDC^11^ the case fatality rate (CFR) was 0.2%: 2 deaths in 1036 confirmed cases on the last day of January 2020. Yet, 14.4% of the cases were considered severe or even critical.

As stated on the page Coronavirus COVID-19 Global Cases by Johns Hopkins CSSE^12^ there were 79441 cases confirmed worldwide (97% on mainland China) and the number of deaths is 2620, 95% of which have occurred in mainland China, at 10 a.m. (Greenwich time zone) of the 24th of January. The World Health Organization considers this outbreak a “public health emergency of international concern”^13^.

The epidemiological curves of COVID-19 in China^11^ are thought to be of extreme value, presenting the progression of illness in the outbreak over time from December 8, 2019 to February 11, 2020, when there were a total of 72314 confirmed cases as the geo-temporal spread of COVID-19 presented in figure 2 of the same paper^11^ to establish the epidemiological state of COVID-19 in China. Moreover, the authors presented age, gender and comorbidity case fatality rate in China in an univariate way.

**Figure 1.**
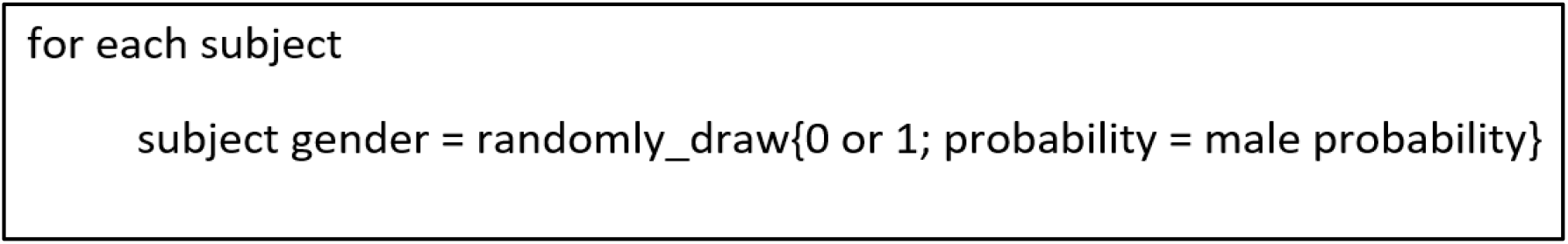
Pseudocode describing how gender variable is created in the dataset. Female is 0; male is 1.

**Figure 2.**
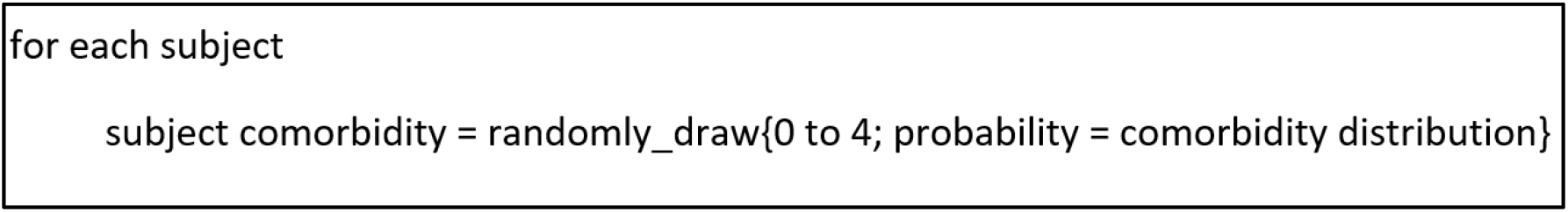
Pseudocode describing how comorbidity variable is created in the dataset. None is 0, hypertension is 1, diabetes is 2, cardiovascular is 3, respiratory is 4 and cancer is 5.

Thus, our main goal is to establish the odds ratio for COVID-19 mortality, adjusted to age and gender-specific prevalence of the morbidities reported in the CDC paper^11^, namely hypertension, diabetes, cardiovascular disease, chronic respiratory disease and cancer.

The risk factors for MERS-CoV were established after analysing data collected from the World Health organization between 2012 and 2018^14^, where males seemed to be more affected than females, and also age over 30 years-old or the presence or comorbidities. Besides these baseline characteristics, ethnicity also presented statistically significant odds ratios. The search performed in Pubmed on the 21^th^ of January 2020 using the following keywords ((“odds ratio” OR “risk ratio” OR “relative risk” OR risk)) AND (SARS-CoV OR SARS OR MERS-CoV OR MERS OR corona OR coronavirus) and updated on the 24^th^ of January 2020 produced no results for studies reporting risk factors for SARS-CoV. Therefore, it is of extreme importance to briskly define the odds ratio for mortality adjusted to comorbidities which are highly prevalent globally, according to age and gender, in order to prevent COVID-19 specific mortality.

## Material and Methods

The main goal of the present work is to compute adjusted odds ratio (OR) for death from COVID-19 considering age intervals, gender and comorbidities as possible risk factors. Complete data is not accessible but a thorough description of the outbreak was made available by China CDC on the 14^th^ February^11^. Based on these data we implemented a Monte Carlo simulation^§^ in the R programming language^15^ to create a synthetic dataset from which a logistic regression can be computed along with ORs.

The dataset includes four variables: age, gender, comorbidities and death. All variables are treated as qualitative and follow the same rationale expressed in the China CDC report. Age is described by intervals of decades (0-9; 10-19; up to ≥ 80 years old) and the comorbidities are: hypertension, diabetes, cardiovascular disease, chronic respiratory disease and cancer. The outcome variable, death, is dichotomous with states dead and alive.

The aforementioned report has information about age and gender distributions, which we used to populate the dataset. However, these distributions are described independently making it impossible to know the exact gender frequency for each age interval. We assumed that the distribution by gender would be similar across age intervals. To render the gender, male or female, for each subject a random number is drawn following a Bernoulli random experience with probability given by the gender distribution (Figure 1).

A similar procedure is performed to attribute one comorbidity to each subject of the dataset: a random number with five possible outcomes (four for comorbidities and one for no disease) is drawn and the corresponding probability is given by an adequate distribution (Figure 2).

A CSV (Comma-Separated Values) file containing the comorbidity distribution by age and gender feeds the routine. Comorbidities’ empirical prevalence can be computed from China CDC report for all subjects, but does not allow different values for each age interval and gender and, for that reason, we considered that prevalence values for each comorbidity were constant across age groups and gender. Despite this limitation, tests can be run using worldwide prevalence for each disease. Finally, comorbidities classes were assumed to be exclusive, i.e., no subject could have more than one comorbidity.

After populating age, gender and comorbidities variables in the dataset, it still misses the death variable, which highly depends on the previous items. To fill in this variable, we used a Naïve Bayesian approach considering that age, gender and comorbidities would be independent. Therefore, the probability, *P*(*D*|*A, G, C*), of dying given the age interval (*A*), the gender (*G*) and the comorbidity (*C*) is given from

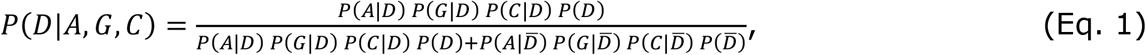

where *D* stands for dying (or death), *G* for gender, *C* for comorbidity and 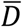 is the complementary of dying, which is to be alive. The notation *P*(*A*|*B*) means probability of A given B. The values at the right-hand side of Equation 1 can be computed from the China CDC report; thus, for each situation we can calculate the probability of being dead and, therefore, to randomly draw a value describing the status of each subject (Figure 3).

**Figure 3.**
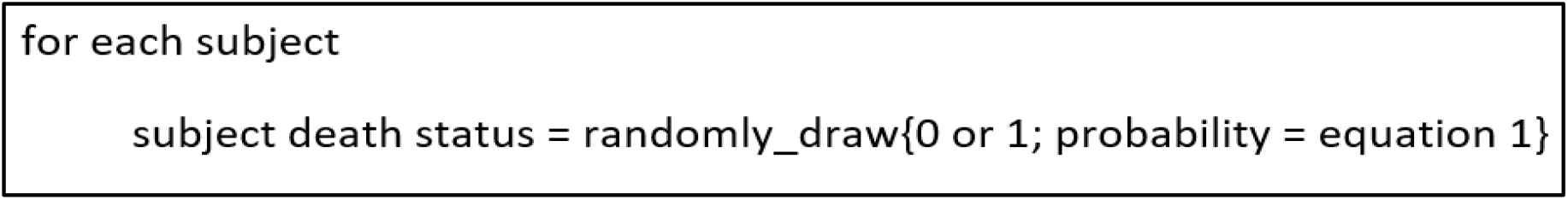
Pseudocode describing how death variable is created in the dataset. Alive is 0 and dead is 1.

Once the dataset is created, a logistic regression model is fitted taking as dependent variable the death variable and as independent variables age, gender and comorbidity. The values of the regression coefficients are then transformed with an exponential function in order to obtain the adjusted OR, which are stored. The procedure is repeated 1000 times, allowing to obtain different datasets, each one with corresponding ORs (Figure 4). After ending the random sampling procedure, the median is computed as well as percentiles 2.5 and 97.5, which allows to obtain the 95% confidence interval of ORs.

**Figure 4.**
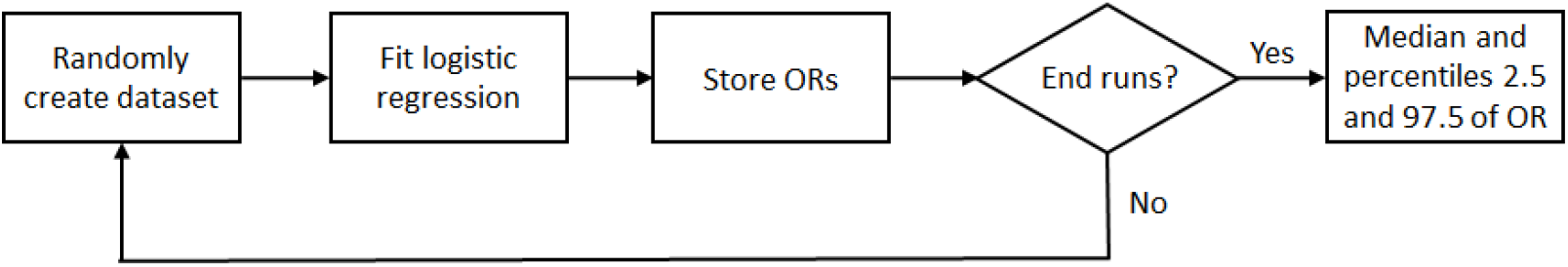
Flowchart of the Monte Carlo routine employed to build synthetic datasets and compute odds ratio.

The validation of the procedure is performed by comparing a set of probability values that are computed both in the synthetic data and in the China CDC report. This set of values includes the gender, death and comorbidities probabilities as well as conditional death probabilities regarding the comorbidities.

## Results

Table 1 shows the statistics regarding the synthetic datasets and the corresponding values from the China CDC report.

**Table 1.** Relative frequencies and corresponding confidence intervals for 1000 simulated datasets and the actual values.

| Statistic | Synthetic (%) | Real# (%) |
| --- | --- | --- |
| Male | 51.43 CI95%[50.98; 51.90] | 51.44 |
| Subjects without comorbidities | 76.83 CI95%[76.52; 77.15] | 74.65 |
| Subjects with hypertension | 12.74 CI95%[12.52; 12.88] | 12.89 |
| Subjects with diabetes | 4.62 CI95%[4.44; 4.82] | 5.30 |
| Subjects with cardiovascular disease | 3.47 CI95%[3.29; 3.64] | 4.20 |
| Subjects with chronic respiratory | 1.95 CI95%[1.82; 2.08] | 2.46 |
| Subjects with cancer | 0.39 CI95%[0.34; 0.45] | 0.51 |
| CFR | 2.20 CI95%[2.07; 2.33] | 2.42 |
| CFR in males subjects | 2.75 CI95%[2.54; 2.96] | 2.84 |
| CFR in subjects without comorbidities | 0.91 CI95%[0.82; 1.01] | 0.86 |
| CFR in subjects with hypertension | 5.74 CI95%[5.19; 6.32] | 6.00 |
| CFR in subjects with diabetes | 6.78 CI95%[5.80; 7.95] | 7.26 |
| CFR in subjects with cardiovascular disease | 8.97 CI95%[7.60; 10.42] | 10.54 |
| CFR in subjects with chronic respiratory disease | 5.97 CI95%[4.41; 7.67] | 6.26 |
| CFR in subjects with cancer | 5.37 CI95%[2.41; 8.77] | 5.61 |

#values computed from CDC China report<sup>11</sup>

Slight departures are observed in some of the frequencies (e.g. comorbidities and CFR) but in general, the reported frequencies are within the confidence intervals meaning that synthetic datasets are reasonably representative of the observed data. Still, this is a limitation of the method.

Table 2 shows the adjusted odds ratio resulting from the logistic regression model and their corresponding confidence intervals. The confidence intervals were obtained by considering as its limits the percentiles 2.5 and 97.5 of the OR list. At each run, only ORs that were statistically significant (p < 0.05; except for age interval ‘40-49’ where p < 0.10 was used) were considered for the final list, hence the actual number of iterations can differ for each OR, which is shown in the last column (# runs) of the table.

**Table 2.** Adjusted ORs for age intervals, gender and comorbidities.

| Risk factor | Adjusted OR | # runs |
| --- | --- | --- |
| Age: 20-29 years old | 0.2017 CI95%[0.0862; 0.3373] | 27 |
| Age: 30-39 years old | 0.3271 CI95%[0.2247; 0.3970] | 19 |
| Age: 40-49 years old | 5.6030 CI95%[0.6917; 6.5224] | 44<sup>§</sup> |
| Age: 50-59 years old | 6.7626 CI95%[2.8942; 15.1582] | 811 |
| Age: 60-69 years old | 18.8161 CI95%[7.1997; 41.5517] | 837 |
| Age: 70-79 years old | 43.7291 CI95%[16.4683; 96.9692] | 837 |
| Age: ≥80 years old | 86.8680 CI95%[32.5717; 202.4672] | 837 |
| Sex: male | 1.8518 CI95%[1.5996; 2.1270] | 1000 |
| Comorbidity: hypertension | 7.4191 CI95%[6.3281; 8.7941] | 1000 |
| Comorbidity: diabetes | 9.0329 CI95%[7.3926; 11.3479] | 1000 |
| Comorbidity: cardiac disease | 12.8328 CI95%[10.2736; 15.8643] | 1000 |
| Comorbidity: chronic respiratory disease | 7.7925 CI95%[5.5446; 10.4319] | 1000 |
| Comorbidity: cancer (any) | 6.8845 CI95%[3.3539; 12.4529] | 1000 |

<sup>§</sup>Obtained with $\alpha = 0.10$ . Does not reach statistical significance

Logistic models assume a reference class for each qualitative independent variable and the ORs interpretation of the remaining classes is made relatively to the reference. In the present case, the reference classes were: ‘0-19’ years old, ‘female’ and ‘no comorbidities’. Therefore, the OR=1.8518 for males means that the ratio of the odd of dying to the odd of not dying is 1.8518 times greater in males than in females. As expected ORs increase with age, but the rate of increasing is very high. ORs for comorbidities were expected to be greater than 1, but once again the values are somehow greater than what one could have anticipated.

## Discussion and conclusion

The objective of this work was accomplished as adjusted ORs were obtained, which allows a direct comparison between risk factors and the assessment of their importance for the final outcome (dying from COVID-19). Analyzing the ORs, it can be learnt that elderly patients (above 60 years old) present the highest risk (OR=18.8161 CI95%[7.1997; 41.5517]) to COVID-19, even greater than having any comorbidity. On the other hand, it seems that younger adult patients (30-39 years old) present some protection (OR=0.3271 CI95%[0.2247; 0.3970]). Regarding comorbidities, cardiovascular disease appears to be the riskiest (OR=12.8328 CI95%[10.2736; 15.8643], which is higher than having a chronic respiratory disease (OR=7.7925 CI95%[5.5446; 10.4319]), but without statistical significance since the confidence intervals overlap. In what concerns gender, males are at risk with OR=1.8518 (CI95%[1.5996; 2.1270]).

To have a point of comparison, patients aged more than 30 years old and affected by MERS-CoV presented OR=2.38 (CI95%[1.75; 3.22])^14^. And MERS-CoV patients with comorbidities had OR=1.76 (CI95%[1.39; 2.22])^14^.

The approach adopted in this work has several limitations that must be referred to put into perspective the results. The first aspect, is that case fatality rate (CFR) is a feeble measure as it highly depends on the correct number of diagnosed cases and also on the final outcome of the current cases. Since this is an in-progress situation, CFR value is naturally uncertain, which undermines all parameters computed from this value, and naturally, the results obtained herein suffer from this fact.

Regarding the CFR value, another question that arose was that the China CDC report enables two ways to compute it. All the observations may be used (1023 deaths in 44672 confirmed cases) or the ones with comorbidities information may be used instead (504 deaths in 20812 confirmed cases). We opted for the latter.

Another aspect is the fact that there is neither information about the correct prevalence of gender per age nor about comorbidities per age and gender. To overcome this issue, we assumed homogeneous prevalence, which will increase ORs in some classes while decreasing in others. This approximation has a non-linear impact difficult to foresee, albeit departures from the real case are expected to be small. Moreover, the R routine is prepared to work with complete prevalence information and any user can test it.

We have also assumed independence between risk factors but the logistic regression assumes the same. Therefore, the impact is not expected to be significant.

Concerning the ability to fit a suitable logistic model, we observed that in some cases the model did not present high values for goodness-of-fit tests (data not shown) and some risk factors did not reach statistical significance. We discarded the OR values in these cases, which reduced the number of runs (‘# runs’ column in table 2) for some risk factors. Consequently, the confidence in those values is smaller.

Finally, we ran a simulation as a means to estimate actual values, since the available data is aggregated in groups. The comparison suggests that the differences between simulated and reported data are acceptable. The simulation included several runs intended to surpass this limitation. Confidence intervals were a way of showing the uncertainty associated with the method.

A final point of utmost importance: Equation 1 can be used to determine the probability of dying from COVID-19 given the age interval, the gender and comorbidity of a particular patient. The objective and quantitative risk measurement of each patient or population is paramount to help managing patients segments in health facilities as well as to assist in the determination of public health policies.

## Data Availability

Data used in this work is public.

https://apps.uc.pt/mypage/faculty/fcaramelo/en/covid

## Support

Funded by National Funds via FCT (Foundation for Science and Technology) through the Strategic Project UIDB/04539/2020 and UIDP/04539/2020 (CIBB).

## Footnotes

§ The routine is free available at https://apps.uc.pt/mypage/faculty/fcaramelo/en/covid

## Notes

### Competing Interest Statement

The authors have declared no competing interest.

